# Imaging solute transportation along the posterior lymphatic pathway in the ocular glymphatic system in healthy human participants

**DOI:** 10.64898/2026.04.03.26349283

**Authors:** Xuehua Wen, Yuanqi Sun, Xinyi Zhou, Yinghao Li, Adrian Paez, Jacob Varghese, Jay J. Pillai, Linda Knutsson, Peter C.M. Van Zijl, Richard Leigh, David O. Kamson, Christina R. Graley, Shiv Saidha, Arnold Bakker, Bryan K. Ward, Amir H. Kashani, Jun Hua

## Abstract

**Background:** Recently, a posterior pathway for fluid drainage from the retina to the meningeal lymphatics in the optic nerve (ON) sheath was identified in rodents using intravitreal imaging tracers directly injected into the ocular-globe. Fluid and solute clearance along this pathway may be associated with many diseases. However, intravitreal tracers are rarely used in clinical imaging. As intravenous Gadolinium-based-contrast-agent (GBCA) can enter the globe via the blood-ocular-barriers, it may provide an alternative approach to image this pathway.

**Purpose:** To establish a clinically feasible intravenous GBCA-based MRI approach for tracking fluid and solute transport along the posterior lymphatic pathway in the ocular glymphatic system.

**Materials&Methods:** This prospective study was conducted from March 2021 to September 2022 in healthy participants. Dynamic-susceptibility-contrast-in-the-CSF (cDSC) MRI was performed before, immediately and 4 hours after intravenous-GBCA administration to track GBCA distribution in aqueous humor (AH) and cerebrospinal fluid (CSF) in regions-of-interest (ROIs) in the globe (anterior-cavity, vitreous-body), in the intraorbital and extraorbital ON, and in the intracranial CSF space proximal to the ON (chiasmatic-cistern, interpeduncular-cistern). Kruskal–Wallis tests with post-hoc Dunn’s tests were used for group comparisons.

**Results:** Sixteen healthy participants (mean age±SD: 51±21 years, 5 men) were recruited. Intravenous-GBCA enhancement was observed in all ROIs immediately after injection. At 4-hour-post-GBCA, the vitreous body showed a trend of smaller enhancement area (55±11% versus 49±11%, *P*=.14) and lower GBCA-concentration (0.044±0.014 versus 0.028±0.010 mmol/L, *P*=.07) compared to immediate-post-GBCA. The intraorbital ON showed more widespread enhancement (39±5% versus 59±6%, *P*=.01) and significantly higher GBCA-concentration (0.023±0.009 versus 0.059±0.015 mmol/L, *P*<.001) at 4-hour-post-GBCA.

**Conclusion:** Dynamic fluid and solute transportation along the posterior lymphatic pathway in the ocular glymphatic system in healthy participants was measured by tracking intravenous-GBCAs entering the globe via the blood-ocular-barriers using cDSC-MRI.

**Summary Statement:** Dynamic fluid and solute transportation along the posterior lymphatic pathway in the ocular glymphatic system in healthy participants was measured by tracking intravenous-Gadolinium-based-contrast-agents entering ocular-globe via blood-ocular-barriers using dynamic-susceptibility-contrast-in-the-CSF (cDSC)-MRI.

**Key Results:**

1. In this prospective study of sixteen participants, Gadolinium-based-contrast-agent (GBCA)-induced signal changes were detected in the aqueous-humor and cerebrospinal fluid immediately following intravenous administration using dynamic-susceptibility-contrast-in-the-CSF (cDSC)-MRI.
2. At 4-hour-post-GBCA, the vitreous-body showed a trend of smaller enhancement area (55±11% versus 49±11%, *P*=.14) and lower GBCA-concentration (0.044±0.014 versus 0.028±0.010 mmol/L, *P*=.07) compared to immediate-post-GBCA.
3. The intraorbital-optic-nerve showed more widespread enhancement (39±5% versus 59±6%, *P*=.01) and higher GBCA-concentration (0.023±0.009 versus 0.059±0.015 mmol/L, *P*<.001) at 4-hour-post-GBCA.

## Introduction

The eye is an extension of the central nervous system as they share the same embryological origin. The retina in the eye has neurons and glial cells with similar cellular and molecular characteristics to those in the brain. The optic nerve (ON, cranial nerve II) carries visual information from the retina to the brain(1). Just like the brain, the ON is enclosed by meningeal layers including the dura mater and subarachnoid space (SAS), which contains cerebrospinal fluid (CSF) that is connected with the intracranial CSF space. The ocular-globe contains the aqueous humor (AH), a CSF-resembling fluid produced by the ciliary body and flows into the nearby anterior cavity (AC). Similar to CSF, the AH is essential for waste removal from the eye and for maintaining intraocular pressure. Extensive studies have examined the anterior pathways for AH clearance from the eye via the Schlemm’s canal and the uveoscleral route near AC(1). More recently, a posterior pathway for fluid drainage from the retina to the meningeal lymphatics in the ON sheath driven by the ocular-cranial pressure gradient has been identified (**Figure 1**)(2–6). As the retina has a high metabolic demand, this posterior pathway, referred to as the ocular glymphatic system, may play a crucial role in metabolic waste clearance from the retina in analogy to the glymphatic system in the brain. Impaired solute clearance via the ocular glymphatic system(7, 8) has been associated with various eye diseases such as glaucoma(2, 3). Moreover, many brain diseases such as Alzheimer’s disease and stroke also affect the eye(3, 7, 9, 10). Therefore, the ability to image solute transportation along the posterior lymphatic pathway in the ocular glymphatic system may provide a useful diagnostic tool for a broad range of disorders.

**Figure 1:**
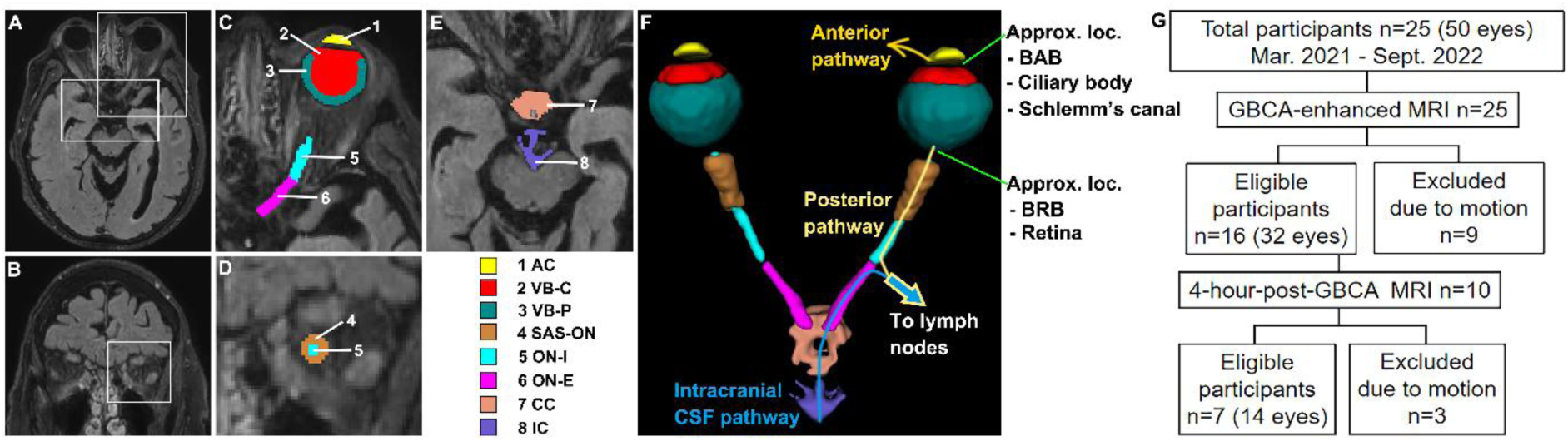
Study overview. **(A,B)** Representative post-GBCA FLAIR MRI images in axial view (z = 87) and in coronal view (y = 236), respectively, from one subject (female). The areas in the white boxes are magnified in subsequent panels. **(C,D,E)** The manually segmented regions of interest (ROIs) are overlaid on the post-GBCA FLAIR MRI images. **List of ROIs**: 1 - AC = Anterior cavity; 2 - VB-C = Central vitreous body; 3 - VB-P = Posterior vitreous body; 4 - SAS-ON = Subarachnoid space around the optic nerve; 5 - ON-I = Intraorbital optic nerve; 6 - ON-E = Extraorbital optic nerve; 7 - CC = Chiasmatic cistern; 8 - IC = Interpeduncular cistern. **(F)** A 3D rendering of the delineated ROIs generated by the software ITK-SNAP. Gadolinium-based contrast agents (GBCA) can enter the eyeball via the blood-aqueous barrier (BAB) in the ciliary body and the blood-retinal barrier (BRB) in the retina. The aqueous humor (AH) in the eye can drain along the conventional anterior pathways through the Schlemm’s canal and the uveoscleral route to the venous and lymphatic systems. More recently, a posterior lymphatic pathway in the ocular glymphatic system from the retina to the optic nerve was identified. The current study aims to develop an intravenous-GBCA-based MRI approach to image this posterior pathway in healthy participants. Intravenous-GBCA can also travel from the intracranial CSF space towards to optic nerve before draining into the lymph nodes. **(G)** Flow diagram showing the number of study participants. Participants with excessive eye movement during MRI were excluded. GBCA: Gadolinium-based contrast agent; FLAIR: fluid attenuated inversion recovery; ROI: region of interest; AH: aqueous humor; CSF: cerebrospinal fluid; BAB: blood-aqueous barrier; BRB: blood-retinal barrier; approx. loc.: approximate location.

The ocular glymphatic system was first identified in rodents using intravitreal imaging tracers directly injected into the vitreous body (VB) in the eyeball(2–4, 6). However, intravitreal tracers are not widely used in humans, mainly due to their invasiveness. One commonly used clinical imaging modality for *in vivo* tracer tracking is Gadolinium-based contrast agent (GBCA)-enhanced MRI, in which GBCA is injected intravenously (IV) to measure blood circulation and perfusion. In healthy human brain, the blood-brain barrier (BBB) is generally impermeable to IV-GBCA, preventing the contrast from entering the perivascular space. Nevertheless, several relatively loose barriers(11) such as the blood-CSF barrier (BCSFB) in the choroid plexus(12) permit IV-GBCAs to cross from blood to the intracranial CSF space. There are two such barriers in the eye: the blood-aqueous barrier (BAB) in the ciliary and iris near the AC; and the blood-retinal barrier (BRB) in the retina. The BAB and BRB are collectively referred to as the blood-ocular barrier (BOB). Numerous reports have documented IV-GBCA accumulation in the VB on post-GBCA clinical MR images in patients with various neurological(10, 13–16) and ophthalmological(17–19) diseases. Furthermore, several studies in healthy participants also observed IV-GBCA enhancement in the eye(20, 21). Therefore, IV-GBCA may provide an alternative approach to direct imaging tracers into the eyeball in a similar way to intravitreal injection, from which the anterior and posterior pathways for fluid clearance from the eye can be imaged.

This technical development sought to establish an IV-GBCA-enhanced MRI approach to image fluid and solute transportation along the posterior lymphatic pathway in the ocular glymphatic system in human participants. As IV-GBCA can present in both blood and CSF/AH, it is critical to isolate GBCA-induced MR signal changes in CSF/AH from blood. Most existing sequences for GBCA-enhanced MRI are sensitive to signal changes from both blood and CSF/AH, resulting in substantial partial volume effects. Recently, the dynamic-susceptibility-contrast-in-the-CSF (cDSC) MRI approach(22) was developed, which uses a long echo time (TE) to effectively suppress blood signals, allowing for selective detection of CSF signals with minimal partial volume effects. Using cDSC-MRI, GBCA-induced CSF signal changes can be robustly measured around highly vascularized brain regions such as the choroid plexus(12) and olfactory areas(23). Technical details about cDSC-MRI are described in the **Supplementary Materials**. Here, by performing cDSC-MRI immediately and 4 hours following IV administration, we examine the entrance points at the BAB and BRB for IV-GBCA to access the eyeball and track the transportation of GBCA in the fluid along the posterior lymphatic pathway in the ocular glymphatic system in a group of healthy participants.

## Methods

### Study design

This prospective study was conducted from March 2021 to September 2022 in healthy participants (**Figure 1**). It was approved by the Institutional Review Board, and written informed consent was obtained from every participant. Exclusion criteria included prior ophthalmologic, neurological, or psychiatric disorders; inability to perform MRI, and excessive eye movement during MRI. All participants completed the immediate-post-GBCA MRI scans, and a subset completed the 4-hour-delayed post-GBCA scans. Participants were selected randomly for technical development.

### MRI

All MRI scans were performed using a 3T scanner (Philips Healthcare, Best, The Netherlands). A 32-channel phased-array head coil was used for signal reception and a dual-channel body coil for transmission. The GBCA Prohance (0.5 mmol/mL; Bracco S.p.A., Milan, Italy) was administered intravenously following standard protocol (dosage: 0.1 mmol/kg, injection rate: 5 mL/s) during MRI.

All participants completed the following MRI scans:

a. Pre-GBCA Fluid-attenuated-inversion-recovery (FLAIR) MRI: repetition-time (TR)/inversion-time (TI)/TE=6000/2000/180ms, voxel=0.8x0.8x0.8mm^3^, 188 slices, three-dimensional turbo-spin-echo readout, 8 minutes 36 seconds.
b. Dynamic cDSC MRI performed continuously before and after GBCA administration: TR/TE/echo spacing (ES)=10000/1347/3.2ms, voxel=0.8x0.8x0.8mm^3^, 188 slices, same coverage as FLAIR, three-dimensional turbo-spin-echo readout. The total duration was 10 minutes 20 seconds including 28 pre-GBCA and 32 post-GBCA volumes with a temporal resolution of TR=10s.
c. Post-GBCA FLAIR MRI: identical parameters as (a).

The 4-hour follow-up MRI session included the same FLAIR and cDSC MRI scans.

### Data analysis

The Statistical-Parametric-Mapping (SPM) software (London, UK) and Matlab (MathWorks, Natick, MA, USA) were used for image analysis. Head motion during dynamic cDSC MRI scans was corrected using the realignment routine in SPM. After that, the mean cDSC image was used as the reference for co-registration with the FLAIR images. Eight regions-of-interest (ROIs) in the ocular areas were manually segmented on post-GBCA FLAIR images using the ITK-SNAP software (Version 4.2.0) with the co-registered cDSC images as a secondary reference (**Figure 1**):

**1-3)** Regions in the eyeball: anterior cavity (AC), central (VB-C) and posterior vitreous body (VB-P). The AC and VB are separated by the lens and the zonular fibers (suspensory ligaments). The BAB is in the ciliary body and iris around the posterior boundary of AC. The VB is further divided into the central (VB-C) and posterior (VB-P) parts with a boundary located approximately 3 mm inside the ocular wall. The VB-P is adjacent to the retina where the BRB is.
**4-6)** Regions in the optic nerve (ON): subarachnoid space (SAS) around the optic nerve (SAS-ON), intraorbital optic nerve (ON-I), and extraorbital optic nerve (ON-E). The SAS-ON was visible mainly around ON-I, which shows hypointensities on FLAIR and hyperintensities on cDSC, as CSF signals are suppressed on FLAIR but enhanced on cDSC with the long TE.
**7,8)** Regions in the intracranial CSF space posterior to the ON: chiasmatic cistern (CC) and interpeduncular cistern (IC). They are separated by the Liliequist membrane, which extends from the posterior clinoid process and dorsum sellae in a posterior-superior direction, with its diencephalic leaf continuing toward and attaching just anterior to the mammillary body, thereby delineating the boundary between CC and IC(24). The CC is connected with the SAS around ON, and both CC and IC are continuous with the intracranial SAS space.

The segmentation was performed by two experienced neuroradiologists (X.W., 12 years’ experience in neuroradiology; and J.J.P., 28 years’ experience in neuroradiology), and all inter-rater discrepancies were resolved through consensus review.

Voxel-wise cDSC relative signal change (ΔS/S) was computed by dividing the difference signal between post- and pre-GBCA with the pre-GBCA signal. To account for potential inter-session signal drift between immediate and 4-hour post-GBCA, all cDSC signals were normalized to the mean corpus callosum signal from the same scan(12). ΔS/S in each ROI was averaged without voxel-selection. GBCA concentration ([Gd]) in CSF was estimated from cDSC ΔS/S following established theories(22) (**Supplementary Material**). Subsequently, voxels exhibiting significant enhancement in each ROI were identified using a one-tailed two-sample t-test comparing post- and pre-GBCA cDSC signals (adjusted *P*<.05). The enhancement area was measured by the percentage of significant voxels in each ROI.

### Statistics

Statistical analysis was conducted using Matlab by X.W.. *P* <.05 was used for significance. Kruskal–Wallis tests with post-hoc Dunn’s tests were used for group comparisons. The Pearson-correlation-coefficient (r) was calculated and multiple regression performed to evaluate the association between GBCA accumulation in the ON and various areas in the ocular-globe and intracranial CSF space. Multiple comparisons were corrected using false-discovery-rate (*P* <.05).

## Results

Sixteen healthy participants (n=32 eyes) were included (mean age±SD: 51±21 years, range: 21-84 years, 5 men) (**Table 1**, **Figure 1**). Among these, seven (n=14 eyes; mean age±SD: 39±17 years, range: 21-69 years; 3 men) underwent 4-hour-post-GBCA MRI. Nine participants with excessive eye movement during MRI were excluded (**Figure 1**). Both eyes from each participant were analyzed separately.

**Table 1:** Demographic characteristics of the study participants.

| Characteristic | Total Participants | GBCA-enhanced<br>MRI | 4-hour-post-GBCA<br>MRI |
| --- | --- | --- | --- |
| No. of participants | 16 | 16 | 7 |
| Age at scan (yr)<sup>†</sup> | 51±21 | 51±21 | 39±17 |
| Sex |  |  |  |
| M | 5 | 5 | 3 |
| F | 11 | 11 | 4 |
| No. of eyes analyzed<sup>‡</sup> | 32 | 32 | 14 |

<sup>†</sup>Data are presented as mean±SD.
<sup>‡</sup>Both eyes from each participant were analyzed.
GBCA: Gadolinium-based contrast agent.

**Table 2** summarizes group results. Immediately following IV-GBCA administration, enhancement was observed in all ROIs in all subjects; among all ROIs, the area near blood-aqueous-barrier (AC) and the area near blood-retinal-barrier (VB-P) showed the highest GBCA concentration ([Gd]) (overall *P*=.02; AC>VB-C: *P*=.03, AC>SAS-ON: *P*=.04, AC>ON-I: *P*=.01, AC>ON-E: *P*=.04, AC>CC: *P*=.04, AC>IC: *P*=.04; VB-P>VB-C: *P*=.08, VB-P>SAS-ON: *P*=.07, VB-P>ON-I: *P*=.03, VB-P>ON-E: *P*=.09, VB-P>CC: *P*=.08, VB-P>IC: *P*=.08), whereas the intraorbital-ON (ON-I) showed the lowest [Gd] (overall *P*=.02; ON-I<AC: *P*=.01, ON-I<VB-C: *P*=.03, ON-I<VB-P: *P*=.02, ON-I<SAS-ON: *P*=.03, ON-I<ON-E: *P*=.04, ON-I<CC: *P*=.03, ON-I<IC: *P*=.04). At 4-hour post-GBCA, the vitreous-body (VB-C, VB-P) showed a trend of smaller enhancement area (VB-C: 58±11% versus 49±12%, *P*=.11; VB-P: 55±11% versus 49±11%, *P*=.14) and lower [Gd] (VB-C: 0.037±0.014 versus 0.034±0.010 mmol/L, *P*=.12; VB-P: 0.044±0.014 versus 0.028±0.010 mmol/L, *P*=.07) compared to immediate-post-GBCA. The enhancement area in AC increased (47±12% versus 59±9%, *P*=.04) at 4-hour-post-GBCA while its [Gd] was comparable to immediate-post-GBCA (0.057±0.020 versus 0.057±0.014 mmol/L, *P*=.46). Along the intraorbital-ON, both SAS-ON and ON-I showed more widespread enhancement (SAS-ON: 44±9% versus 66±6%, *P*=.01; ON-I: 39±5% versus 59±6%, *P*=.01) and significantly higher [Gd] (SAS-ON: 0.039±0.014 versus 0.093±0.020 mmol/L, *P*=.002; ON-I: 0.023±0.009 versus 0.059±0.015 mmol/L, *P*<.001) at 4-hour-post-GBCA. The ON-E, CC, and IC showed comparable enhancement area (ON-E: 56±3% versus 56±5%, *P*=.68; CC: 58±16% versus 60±16%, *P*=.62; IC: 69±15% versus 70±14%, *P*=.58) and slightly higher [Gd] (ON-E: 0.036±0.010 versus 0.042±0.012 mmol/L, *P*=.43; CC: 0.037±0.016 versus 0.052±0.020 mmol/L, *P*=.25; IC: 0.038±0.014 versus 0.041±0.013 mmol/L, *P*=.45) at 4-hour-post-GBCA compared to immediate-post-GBCA.

**Table 2:**
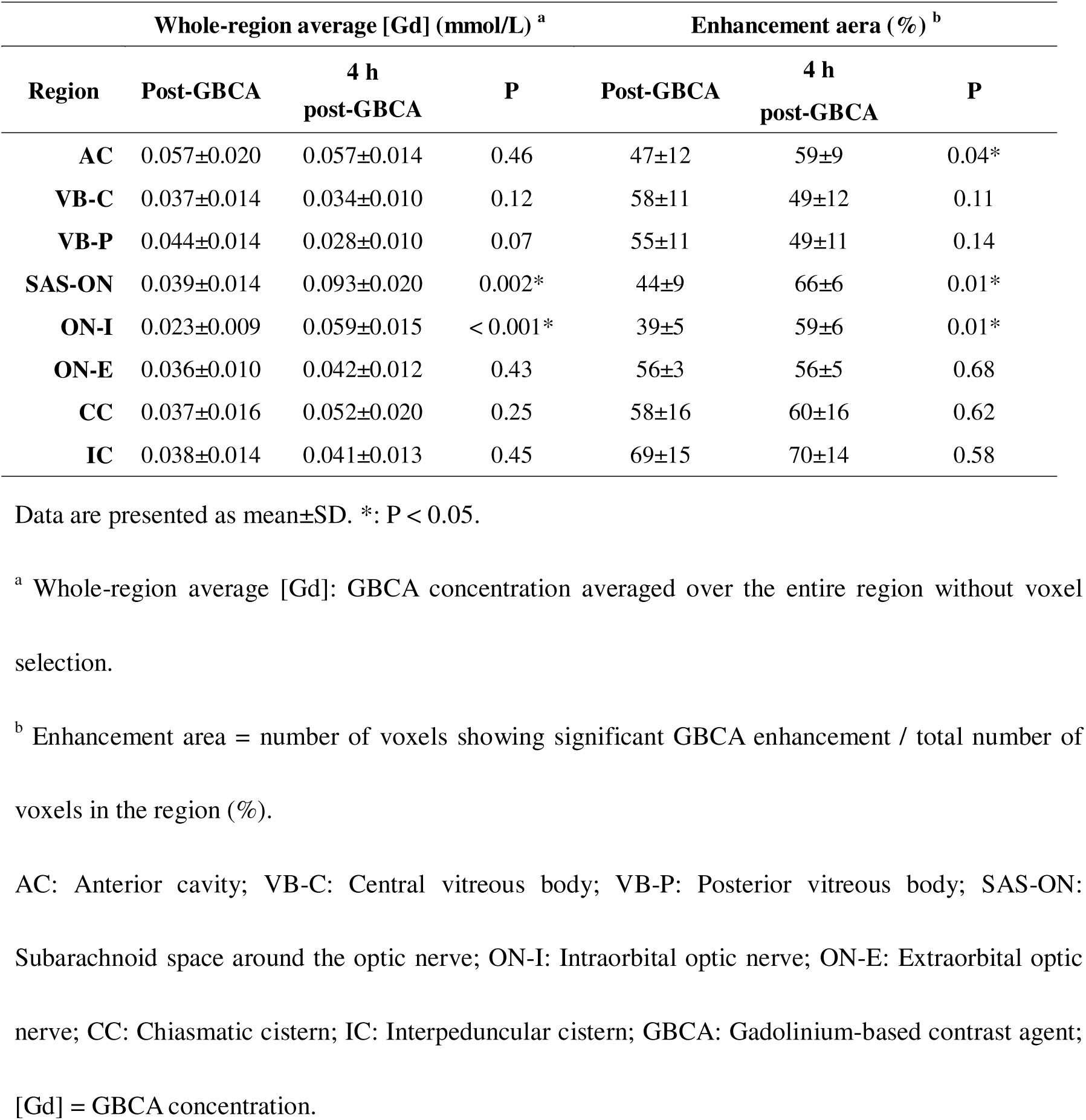
Quantitative results measured from cDSC MRI immediately and 4 hours after intravenous GBCA administration. Significant GBCA enhancement was detected in all regions.

**Figures 2-5** display [Gd] maps in all ROIs overlaid on cDSC images from 4 participants. Compared to group results, individual variations are noted especially in VB-C and VB-P. At 4-hour post-GBCA, the reduced enhancement pattern compared to immediate-post-GBCA in VB-C was most apparent in older subjects (**Figure 2**), but less obvious in younger subjects. However, due to the limited sample size, we cannot conduct a correlation analysis with age.

**Figure 2:**
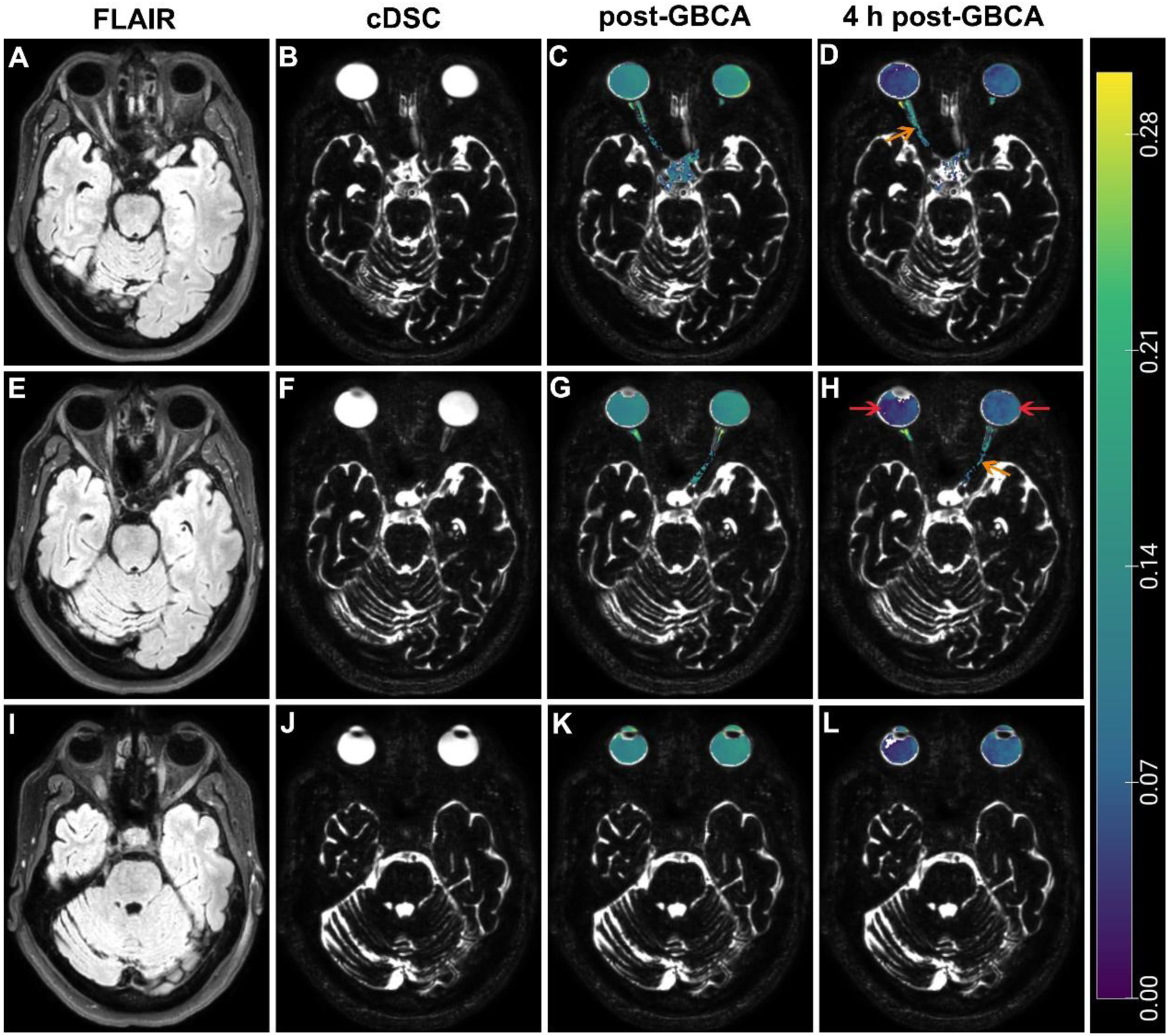
Representative FLAIR **(A, E, I)** and cDSC **(B, F, J)** MRI images and GBCA enhancement maps from a healthy female participant. Three slices in axial view were displayed (first row, **A-D**: z = 70; second row, **E-H**: z = 67; third row, **I-L**: z = 59). GBCA concentration measured from cDSC MRI in the relevant regions immediately **(C, G, K)** and at 4 hours **(D, H, L)** following injection was overlaid on the cDSC images. Red arrow: vitreous body; orange arrow: optic nerve. The color bar indicates GBCA concentration ([Gd]) in mmol/L. GBCA enhancement was observed in the vitreous body immediately following intravenous administration and decreased at 4 hours post-GBCA. GBCA enhancement was observed in the optic nerve immediately following intravenous administration and increased at 4 hours post-GBCA. FLAIR: fluid attenuated inversion recovery; cDSC: dynamic susceptibility contrast in the CSF; CSF: cerebrospinal fluid; GBCA: Gadolinium-based contrast agent.

**Figure 3:**
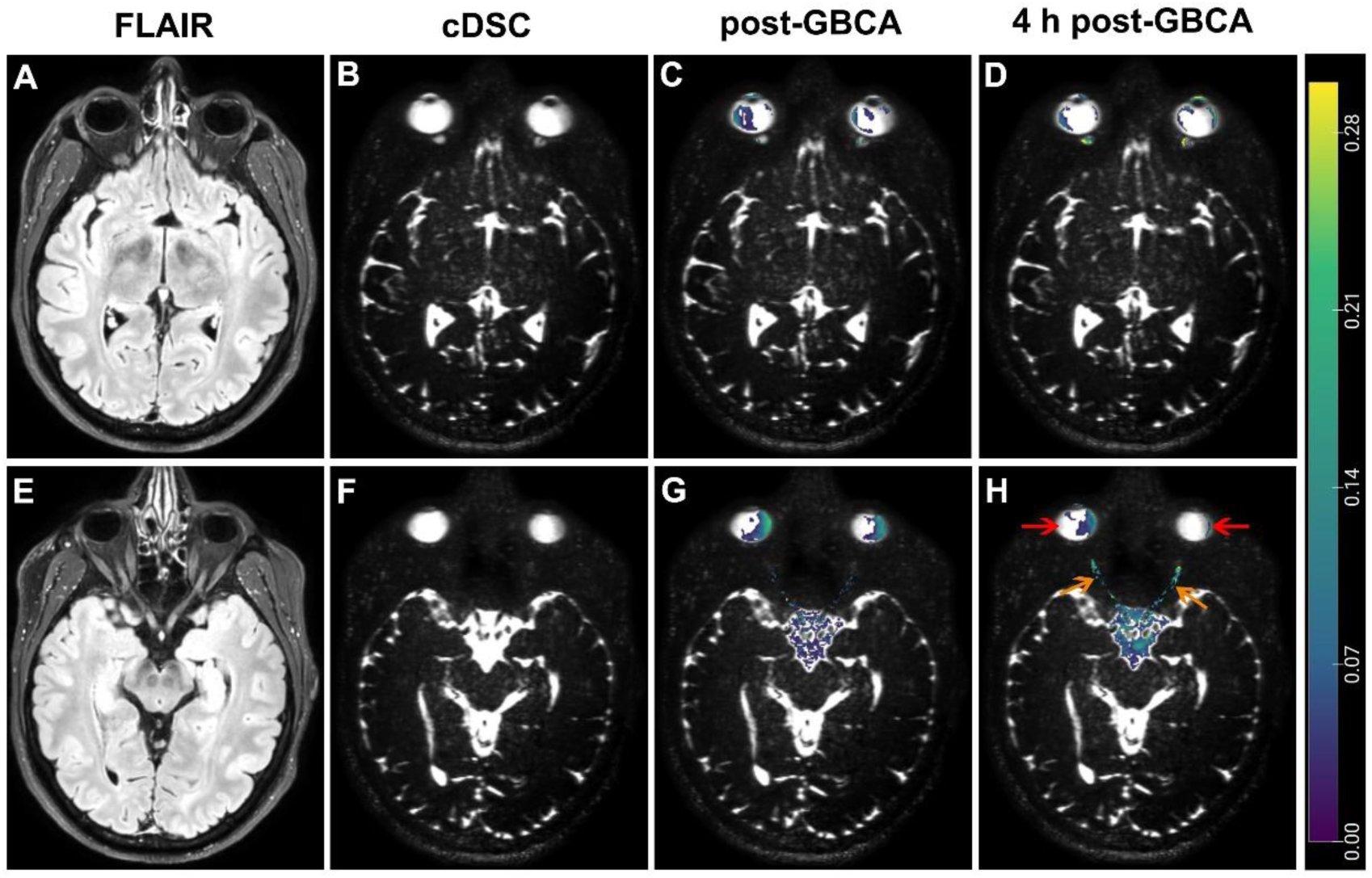
Representative FLAIR **(A, E)** and cDSC **(B, F)** MRI images and GBCA enhancement maps from a healthy male participant. Two slices in axial view were displayed (first row, **A-D**: z = 103; second row, **E-H**: z = 93). GBCA concentration measured from cDSC MRI in the relevant regions immediately **(C, G)** and at 4 hours **(D, H)** following injection was overlaid on the cDSC images. Red arrow: vitreous body; orange arrow: optic nerve. The color bar indicates GBCA concentration ([Gd]) in mmol/L. GBCA enhancement was observed in the vitreous body immediately following intravenous administration and appeared similar at 4 hours post-GBCA. GBCA enhancement was observed in the optic nerve immediately following intravenous administration and increased at 4 hours post-GBCA. FLAIR: fluid attenuated inversion recovery; cDSC: dynamic susceptibility contrast in the CSF; CSF: cerebrospinal fluid; GBCA: Gadolinium-based contrast agent.

**Figure 4:**
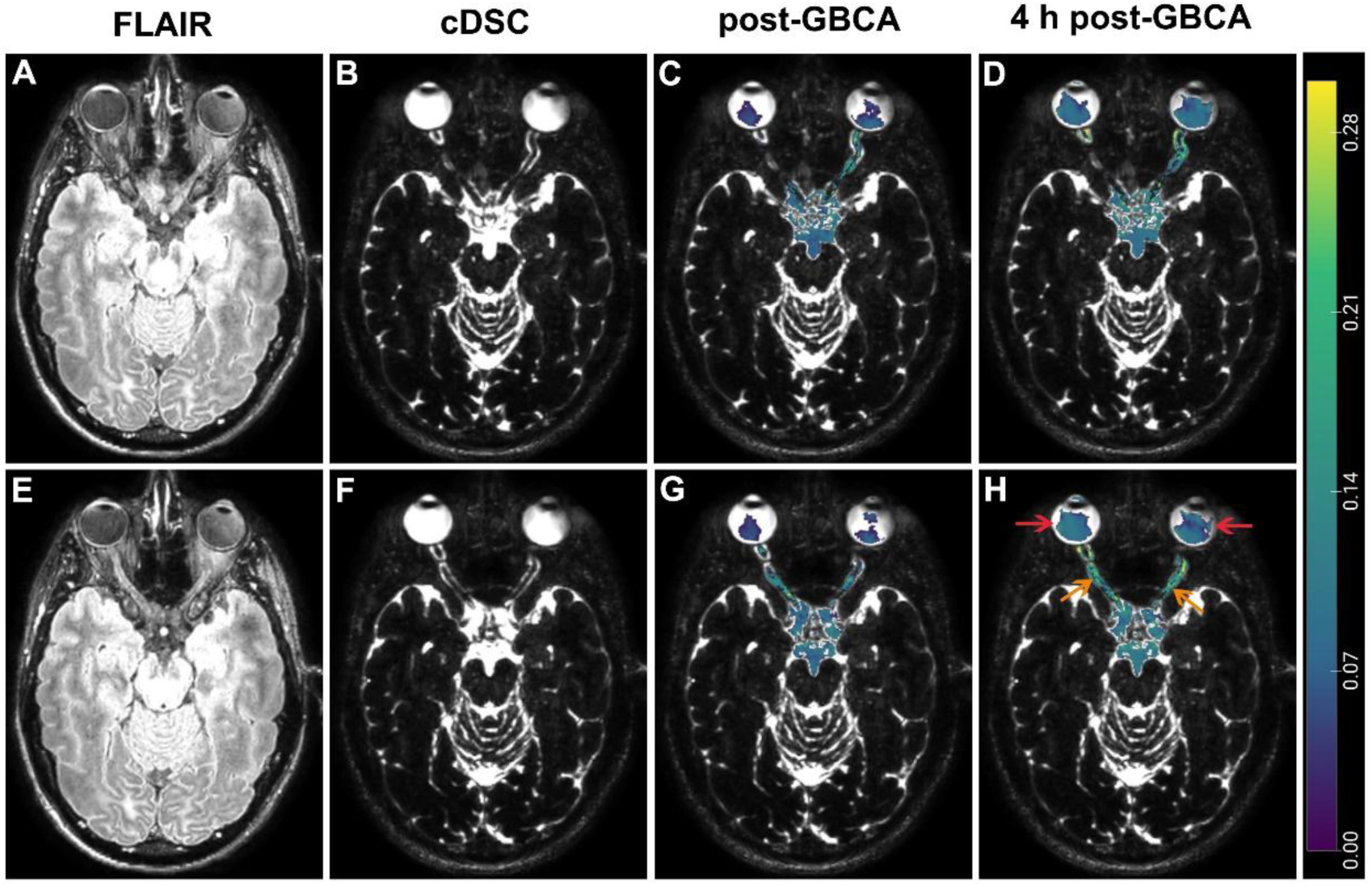
Representative FLAIR **(A, E)** and cDSC **(B, F)** MRI images and GBCA enhancement maps from a healthy female participant. Two slices in axial view were displayed (first row, **A-D**: z = 86; second row, **E-H**: z = 84). GBCA concentration measured from cDSC MRI in the relevant regions immediately **(C, G)** and at 4 hours **(D, H)** following injection was overlaid on the cDSC images. Red arrow: vitreous body; orange arrow: optic nerve. The color bar indicates GBCA concentration ([Gd]) in mmol/L. GBCA enhancement was observed in the vitreous body immediately following intravenous administration and appeared similar at 4 hours post-GBCA. GBCA enhancement was observed in the optic nerve immediately following intravenous administration and increased at 4 hours post-GBCA. FLAIR: fluid attenuated inversion recovery; cDSC: dynamic susceptibility contrast in the CSF; CSF: cerebrospinal fluid; GBCA: Gadolinium-based contrast agent.

**Figure 5:**
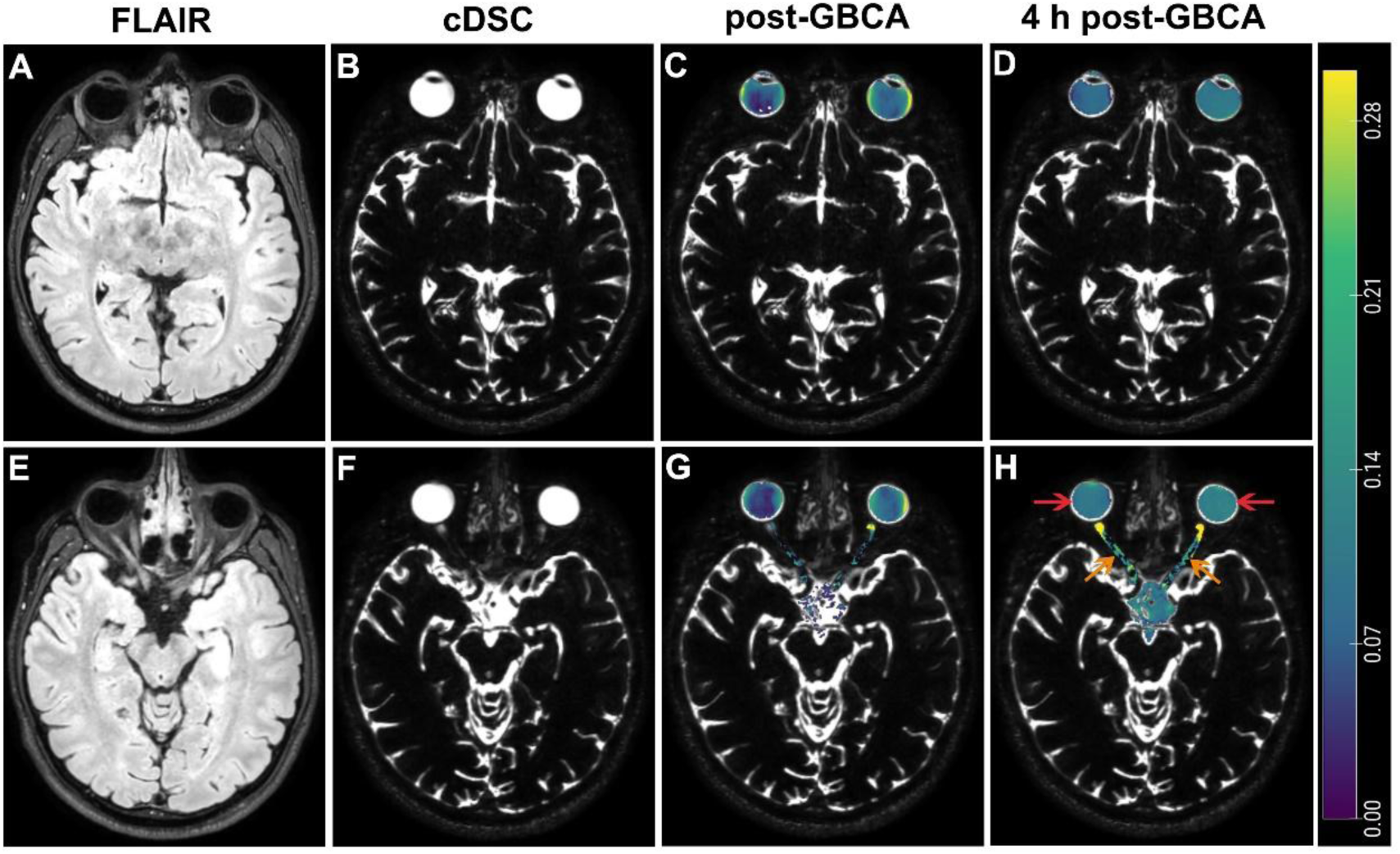
Representative FLAIR **(A, E)** and cDSC **(B, F)** MRI images and GBCA enhancement maps from a healthy male participant. Two slices in axial view were displayed (first row, **A-D**: z = 100; second row, **E-H**: z = 90). GBCA concentration measured from cDSC MRI in the relevant regions immediately **(C, G)** and at 4 hours **(D, H)** following injection was overlaid on the cDSC images. Red arrow: vitreous body; orange arrow: optic nerve. The color bar indicates GBCA concentration ([Gd]) in mmol/L. GBCA enhancement was observed in the vitreous body immediately following intravenous administration and appeared similar at 4 hours post-GBCA. GBCA enhancement was observed in the optic nerve immediately following intravenous administration and increased at 4 hours post-GBCA. FLAIR: fluid attenuated inversion recovery; cDSC: dynamic susceptibility contrast in the CSF; CSF: cerebrospinal fluid; GBCA: Gadolinium-based contrast agent.

**Figure 6** shows correlations of 4-hour-post-GBCA [Gd] between regions. The [Gd] in intraorbital-ON (SAS-ON, ON-I) was significantly correlated with [Gd] in the eyeball regions (AC, VB-C, VB-P) proximal to ON-I (SAS-ON &AC: r=.73, *P*=.005; SAS-ON &VB-C: r=.83, *P*<.001; SAS-ON &VB-P: r=.70, *P*=.007; ON-I&AC: r=.71, *P*=.006; ON-I&VB-C: r=.96, *P*<.001; ON-I&VB-P: r=.86, *P*<.001). The [Gd] in extraorbital-ON (ON-E) was significantly correlated with [Gd] in the intracranial CSF space (CC, IC) directly connected to ON-E (ON-E&CC: r=.81, *P*<.001; ON-E&IC: r=.78, *P*=.002). Multiple regression demonstrated a predominant effect from VB-C on the SAS-ON and ON-I, and from CC on the ON-E (**Table S1**).

**Figure 6:**
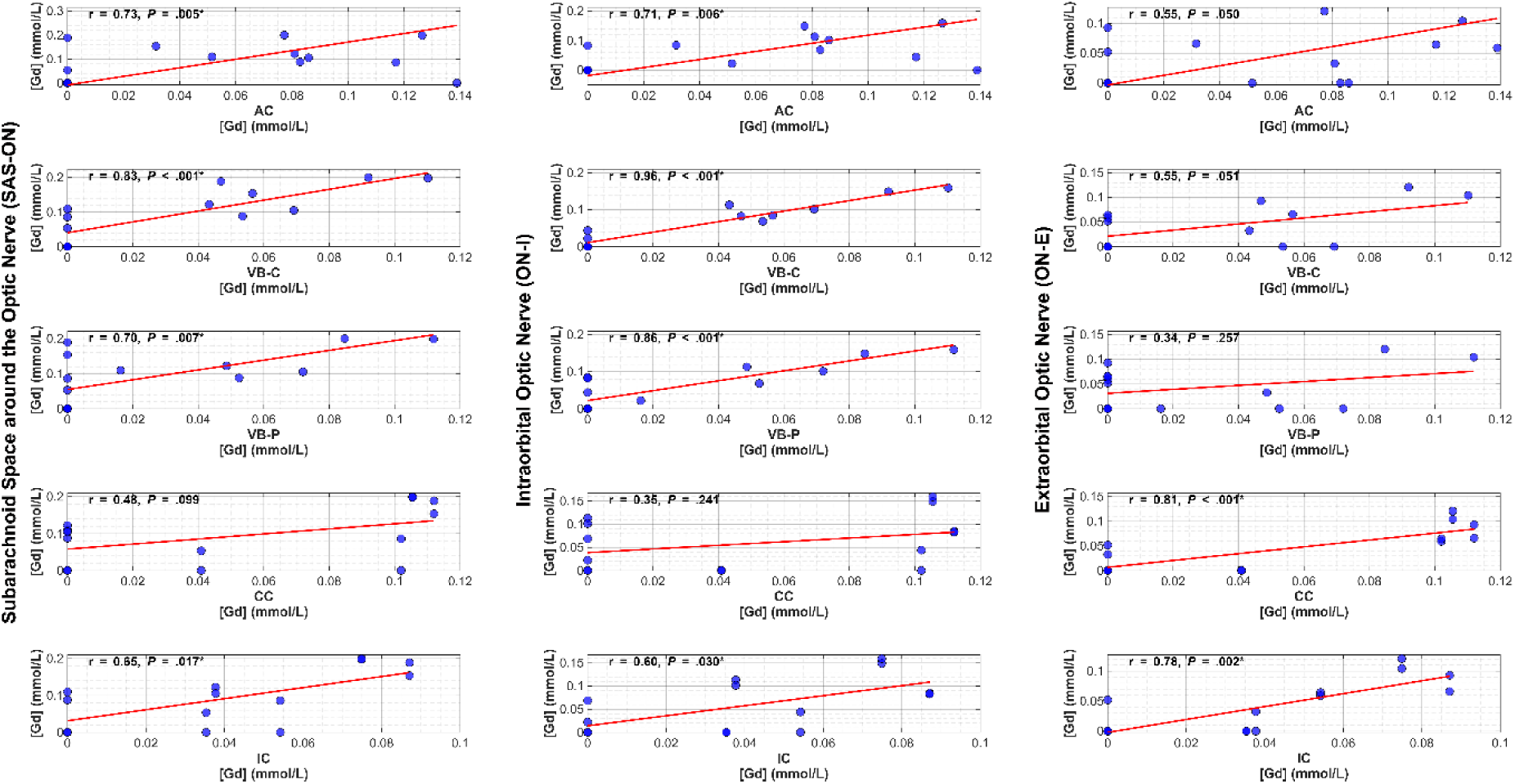
Correlations of GBCA concentrations ([Gd], mmol/L) at 4 hours following intravenous administration between various regions. The subarachnoid space around the optic nerve (SAS-ON), intraorbital optic nerve (ON-I), and extraorbital optic nerve (ON-E) are considered target regions. The anterior cavity (AC), central vitreous body (VB-C), posterior vitreous body(VB-P), chiasmatic cistern (CC), and interpeduncular cistern (IC) are considered potential upstream entry points for GBCAs to reach the ON. Significant correlation was found between AC/VB-C/VB-P and SAS-ON/ON-I, and between CC/IC and ON-E. *: P < 0.05. GBCA: Gadolinium-based contrast agent.

## Discussion

Tracer-based techniques are widely used to visualize fluid and solute transportation. Intravenous (IV)-Gadolinium-based contrast agent (GBCA)-MRI is a commonly performed clinical scan to image blood flow and perfusion. GBCAs can also be injected directly into the cerebrospinal fluid (CSF) via the intrathecal (IT) route, which is often used to image CSF distribution(25). For eye-related regions, IT-GBCA-MRI can track GBCA distribution from intracranial CSF space to the visual pathway(25). However, IT-GBCA-MRI cannot image the recently discovered posterior lymphatic pathway in the ocular glymphatic system from eye to the optic nerve (ON), because the direction of IT-GBCA flow is opposite to the posterior pathway. In rodent studies, intravitreal tracers were used to measure fluid drainage along this posterior ocular pathway(2, 6). In this study, we showed that fluid and solute transportation along this posterior lymphatic pathway can be imaged using IV-GBCA-MRI in healthy human subjects. Immediately following IV administration, GBCA-induced signal changes were detected in the vitreous body (VB) and anterior cavity (AC) using dynamic-susceptibility-contrast-in-the-CSF (cDSC)-MRI, indicating that GBCAs can enter the eyeball via both the BAB near AC and the BRB near retina in healthy participants. Thus, IV-GBCAs can trace fluid clearance pathways from the eye in a manner comparable to intravitreal tracers directly injected into the eyeball. The immediate entrance of IV-GBCAs into the VB aligns with previous reports in patients(10, 13–19) and healthy participants(20, 21). The main difference is that most existing studies on this topic used MRI methods designed to measure GBCA-induced signal changes from blood. A unique feature of this study is that it adopted cDSC-MRI that was optimized to measure GBCA-induced signal changes in the CSF/AH while suppressing blood signals(22). This is particularly important for IV tracers, as it eliminates the ambiguity of signal origin so that the measured cDSC signal changes can be confidently attributed to GBCA distribution in the CSF/AH only, even though the apparent spatial resolution is not enough to visualize small blood vessels and CSF/AH separately. This predominant CSF origin of cDSC MRI signals has been validated in prior technical studies(22).

The intraorbital segment of ON (ON-I) is directly connected with the VB. At 4-hour-post-GBCA, enhancement in the VB decreased, while GBCA enhancement in the intraorbital-ON increased significantly compared to immediate-post-GBCA. This shift of GBCA concentration ([Gd]) indicates a potential posterior drainage pathway from the VB to the ON. These results from IV-GBCA-MRI in humans are consistent with the findings from rodent studies showing fluid and solute drainage from the retina to the meningeal lymphatics in the ON sheath along the posterior pathway revealed by intravitreal tracers(2, 6).

Alternatively, when using IV-GBCAs, the [Gd] increase in the ON may also come from the intracranial CSF space (chiasmatic cistern (CC) and interpeduncular cistern (IC)). The IC and CC showed slightly increased enhancement at 4-hour-post-GBCA. The [Gd] at 4-hour-post-GBCA in IC and CC was comparable to that in ventricular CSF measured in previous IV-GBCA studies in healthy human participants(12), supporting the validity of these measures as IC and CC are connected with ventricular CSF. The extraorbital segment of ON (ON-E) is directly connected with the CC. At 4-hour-post-GBCA, the ON-E showed increased enhancement than immediate-post-GBCA, the magnitude of which was comparable to that of IC and CC, but much lower than that of ON-I. These results from IV-GBCA-MRI agree with IT-GBCA-MRI studies(25), which can image this pathway from intracranial CSF to ON directly.

Taken together, these results imply that GBCAs coming from the intracranial CSF space accumulate primarily in the extraorbital-ON segment adjacent to the CC (ON-E), whereas GBCAs coming from the eye are mainly localized in the intraorbital-ON segment close to the VB (ON-I). This is further supported by the significant correlations of 4-hour-post-GBCA [Gd] between these regions. However, a limitation of this study is that the relative contributions from these two routes cannot be quantified by the proposed method, which warrants further investigation. According to rodent studies, GBCAs reaching the ON will drain through the meningeal lymphatic network in the dura surrounding the ON into lymph nodes eventually(4).

The anterior pathways for AH outflow from the eye via the trabecular meshwork to the Schlemm’s canal and the uveoscleral route were not evaluated in this study. These anterior pathways are considered conventional AH egress pathways which have been studied extensively. The goal of the current study is to establish an IV-GBCA-MRI technique that can be performed on clinical MRI scanners to image the posterior ocular lymphatic pathway.

Our data showed substantial individual variations with age. In older subjects, enhancement in the VB decreased at 4-hour-post-GBCA compared to immediate-post-GBCA. In younger subjects, enhancement in the VB was mostly similar or slightly increased at 4-hour-post-GBCA compared to immediate-post-GBCA. However, as a technical development study, the sample size was very limited. This age correlation requires further evaluation in a larger cohort.

In summary, dynamic fluid and solute transportation along the posterior lymphatic pathway in the ocular glymphatic system in healthy participants was measured by tracking intravenous Gadolinium-based contrast agents entering the eyeball via the blood-ocular-barriers using dynamic-susceptibility-contrast-in-the-CSF (cDSC) MRI. This provides a clinically feasible tool to investigate dysfunction in waste clearance from the eye associated with various ocular and neurological disorders.

## Data Availability

All data produced in the present study are available upon reasonable request to the authors.

## Abbreviations

AH: aqueous humor
AC: anterior cavity
CSF: cerebrospinal fluid
BBB: blood-brain barrier
BCSFB: blood-CSF barrier
BAB: blood-aqueous barrier
BRB: blood-retinal barrier
BOB: blood-ocular barrier
cDSC: dynamic susceptibility contrast in the CSF
FLAIR: fluid attenuated inversion recovery
TR: repetition time
TI: inversion time
TE: echo time
ES: echo spacing
GBCA: Gadolinium-based contrast agent
IV: intravenous
IT: intrathecal
ROI: region of interest
VB: vitreous body
VB-C: central vitreous body
VB-P: posterior vitreous body
ON: optic nerve
SAS: Subarachnoid space
SAS-ON: SAS around the optic nerve
ON-I: intraorbital optic nerve
ON-E: extraorbital optic nerve
CC: chiasmatic cistern
IC: interpeduncular cistern
ΔS/S: relative signal change
[Gd]: GBCA concentration.

## Acknowledgments (anonymized)

The authors thank the staff for their experimental and technical assistance. This work was supported by research grants from national funding agencies.

**No other disclosures to report.**

