## Supplementary file for "Imaging solute transportation along the posterior lymphatic pathway in the ocular glymphatic system in healthy human participants"

**Supplemental Material**

**Brief Description of Dynamic-susceptibility-contrast-in-the-CSF (cDSC) MRI**

The recently developed dynamic-susceptibility-contrast-in-the-CSF (cDSC) MRI approach(1) was adopted in this study to measure Gadolinium-based contrast agent (GBCA)-induced MR signal changes in the CSF. A three-dimensional turbo-spin-echo (TSE, also known as fast spin echo or FSE) MRI sequence with a sub-millimeter spatial resolution, a temporal resolution of ~ 10 seconds and a whole-brain coverage was optimized for cDSC MRI. The key difference between cDSC MRI and many MRI methods adopted in existing studies using GBCA-enhanced MRI is the use of an ultra-long echo time (TE > 600 ms). The T2 relaxation time of CSF (> 1000 ms) is much longer than blood and parenchymal T2 (~ 50-200 ms). A sufficiently long TE can effectively suppress parenchymal and blood signals due to T2 decay. Thus, parenchymal and blood signals in cDSC MRI are in the noise range, not significantly different from the background, and therefore are effectively zero. This furnishes a pure CSF signal with minimal partial volume effects from the blood in cDSC MRI, which is critical for studies on CSF distribution using intravenous GBCAs. The cDSC MRI techniques has been applied in several CSF studies in highly vascularized brain regions such as the choroid plexus(2) and olfactory areas(3). The GBCA-induced MR signal changes in the CSF measured from cDSC MRI has a monotonic relationship with the GBCA concentration in the CSF. This was validated in prior technical development studies by scanning phantoms with various GBCA concentrations(1). Therefore, GBCA concentration in the CSF can be estimated from the cDSC signal changes using the established theory(1). Readers are referred to recent publications for further technical details of the cDSC MRI approach(1-3).

**Table S1.** Multiple regression shows relative contributions from various brain regions to GBCA concentration in the ON measured at 4 hours after intravenous administration. GBCA concentration at 4 hours after injection in VB-C had a dominant effect on that in the intraorbital segment of ON (SAS-ON, ON-I). GBCA concentration at 4 hours after injection in the CC had a dominant effect on that in the extraorbital segment of ON (ON-E).

| | Standardized coefficient | | | | | P | | | | | | |
|---|---|---|---|---|---|---|---|---|---|---|---|---|
| **Region** | **AC (β1)** | **VB-C (β2)** | **VB-P (β3)** | **CC (β4)** | **IC (β5)** | **AC (β1)** | **VB-C (β2)** | **VB-P (β3)** | **CC (β4)** | **IC (β5)** | **$R^2$** | **Adjusted $R^2$** |
| **SAS-ON** | -0.080 | **1.436** | -0.121 | 0.003 | 0.647 | 0.845 | **0.274** | 0.921 | 0.996 | 0.625 | 0.745 | 0.585 |
| **ON-I** | 0.126 | **0.979** | 0.190 | -0.352 | 0.797 | 0.433 | **0.072** | 0.688 | 0.230 | 0.144 | 0.936 | 0.896 |
| **ON-E** | -0.098 | 0.230 | 0.259 | **0.853** | -0.351 | 0.670 | 0.748 | 0.707 | **0.064** | 0.638 | 0.761 | 0.612 |

The following models were used in multiple regression:

[Gd] in SAS-ON/ON-I/ON-E = β0 + β1×[Gd] in AC + β2×[Gd] in VB-C + β3×[Gd] in VB-P + β4×[Gd] in CC + β5×[Gd] in IC

The Standardized coefficients (β1- β5) can be used as an estimate of relative contributions from each term.

AC: Anterior cavity; VB-C: Central vitreous body; VB-P: Posterior vitreous body; SAS-ON: Subarachnoid space around the optic nerve; ON-I: Intraorbital optic nerve; ON-E: Extraorbital optic nerve; CC: Chiasmatic cistern; IC: Interpeduncular cistern; GBCA: Gadolinium-based contrast agent; [Gd] = GBCA concentration.
